# Association of changing physical activity intensity and bout length with mortality: a study of 79,507 participants in UK Biobank

**DOI:** 10.1101/2020.05.22.20108795

**Authors:** Louise AC Millard, Kate Tilling, Tom R Gaunt, David Carslake, Deborah A Lawlor

## Abstract

**Background:** Spending more time active (and less time sedentary) is associated with many health benefits such as improved cardiovascular health and lower risk of all-cause mortality. However, it is unclear whether these associations differ depending on whether time spent sedentary or in moderate-vigorous physical activity (MVPA) is accumulated in long or short bouts. In this study we used a novel analytical approach, that accounts for substitution (i.e. more time in MVPA means spending less time sleeping, in light activity or being sedentary), to examine whether length of sedentary and MVPA bouts associates with all-cause mortality.

**Methods and findings:** We used data on 79,507 participants from UK Biobank. We derived the total time participants spent in activity categories – sleep, sedentary, light activity and MVPA – on average per day. We also derived the time spent in sedentary and MVPA bouts of short (1-15 minutes), medium (16-40 minutes) and long (41+ minutes) duration. We used Cox proportion hazards regression to estimate the association of spending 10 minutes more average daily time in one activity or bout length category, coupled with spending 10 minutes less time in another, with all-cause mortality. Those spending more time sedentary had higher mortality risk if this replaced time spent in light activity (hazard ratio 1.02 [95% confidence interval (CI): 1.01, 1.03]), and an even higher risk if this replaced time spent in MVPA (hazard ratio 1.08 [95% CI: 1.06, 1.10]). Those spending more time in MVPA had lower mortality risk, irrespective of whether this replaced time spent sleeping, sedentary or in light activity. We found little evidence to suggest that mortality risk differed depending on the length of sedentary or MVPA bouts. A limitation of our study is that we cannot assume that these results are causal, though we adjusted for key confounders.

**Conclusions:** Using our novel analytical approach, we uniquely show that time spent in MVPA is associated with reduced mortality, irrespective of whether it replaces time spent sleeping, sedentary or in light activity. This emphasizes the specific importance of MVPA. We found little evidence to suggest that the impact of MVPA differs depending on whether it is obtained from several short bouts or fewer longer bouts, supporting recent policy changes in some countries. Further studies are needed to investigate causality and explore health outcomes beyond mortality.

## INTRODUCTION

Physical activity is associated with many health benefits such as better cardiovascular health, and reduced risk of some cancers and type II diabetes (1). A recent systematic review of prospective studies suggested that higher levels of physical activity at any intensity, and less time spent sedentary, are associated with a reduced risk of mortality (2).

Policy in the UK and US recommends that people accumulate 150 minutes each week in moderate-vigorous physical activity (MVPA), or 75 minutes in vigorous activity (3,4). Until recently, the advice also stated that activity should be accumulated in bouts of 10 minutes or more, but this was changed as a result of evidence from a meta-analysis that included 19 randomised trials and concluded that adults are likely to have similar cardiovascular benefit irrespective of bout duration (5). These trials had small sample sizes (all < 300) and short term follow-up (typically up to 4 weeks long (5)) hence are only able to measure continuously measured risk factors. Furthermore, all of the trials in the meta-analysis were categorised as having a high risk of bias (5).

Few observational studies have assessed how duration of MVPA bouts relates to health. A meta-analysis of 29,734 children (4-18 years old) across 21 cohort studies found a similar benefit of MVPA on cardiometabolic risk factors across different bout durations (6). In that study an isotemporal approach was used to estimate associations of spending more time in one MVPA bout duration category coupled with less time in another MVPA bout duration category. They controlled for the overall time in MVPA to investigate its composition, but did not account for time spent in other activity categories such as sleeping or sedentary (6). Of two studies in adults, one (N=2,190) found no notable association of MVPA bout length with cardiovascular risk factors (7). The other (N= 3,250), reported smaller mean waist circumference and lower BMI in those who spent more time in MVPA bouts of 10+ minutes rather than shorter bouts (8). Both these studies did not consider couplings of activity categories, thus did not examine whether results differed depending on the form of activity substituted for MVPA. All those studies grouped bouts ≥10 mins together, probably because they lacked power to explore more bouts length categories (6–8). Other studies have used two statistics to characterise MVPA bouts: 1) the number of bouts, and 2) the average time spent in bouts (in total) per day, but these do not describe the range of bout lengths a person undertakes or how often they undertake them (9-13). We have found only one study that examined the importance of sedentary bout length (N = 7,985 adults) (14). It found that higher percentage of total sedentary time in shorter sedentary bouts (<=29 mins) was associated with lower mortality, but overall time spent sedentary was not accounted for (Supplementary figure 1).

The aim of our study is to examine whether mortality differs depending on time spent in different activity categories, and whether time spent sedentary or in MVPA is accumulated in longer versus shorter bouts. We use a novel analytical approach that addresses flaws and limitations of previous studies, to assess associations of overall time spent in different activity categories and bout length categories in terms of coupling more time spent in one category with less time in another category. Our isotemporal approach splits complete days into all activity categories – sleep, sedentary, light and MVPA – to estimate associations of, for example, spending more time in long MVPA bouts coupled with less time sedentary. Our study is the largest observational study of activity bouts to date and is the first to assess associations of MVPA bout length with all-cause mortality.

## METHODS

### Participants

We used data from UK Biobank participants. UK Biobank is a large prospective cohort of approximately 500,000 adults (5% of those invited (15)) aged 40-69 years old at recruitment between 2006 and 2010 (16). Written informed consent was obtained to collect and store data and biosamples and link participants to health and administrative data, and for researchers to use these data for health research. UK Biobank received ethical approval from the UK National Health Service’s National Research Ethics Service (ref 11/NW/0382).

In 2013 a subsample of 240,000 UK Biobank participants were randomly selected from those who had provided valid email addresses, and invited to participate in the accelerometer sub-study. Participants of one assessment centre (3797 participants; 0.76% of the cohort) were excluded from being invited as they had been invited to participate in pilot studies (17). Between 2013 and 2015 participants were sent devices in order of acceptance. Of those invited, 106,053 agreed to participate, and 103,711 (44% of invited) provided some accelerometer data (17). Our study includes 84,182 participants with at least 72 hours accelerometer wear time and no missing data for confounding factors used in our analyses (Figure 1 and Supplementary section S1).

**Figure 1:**
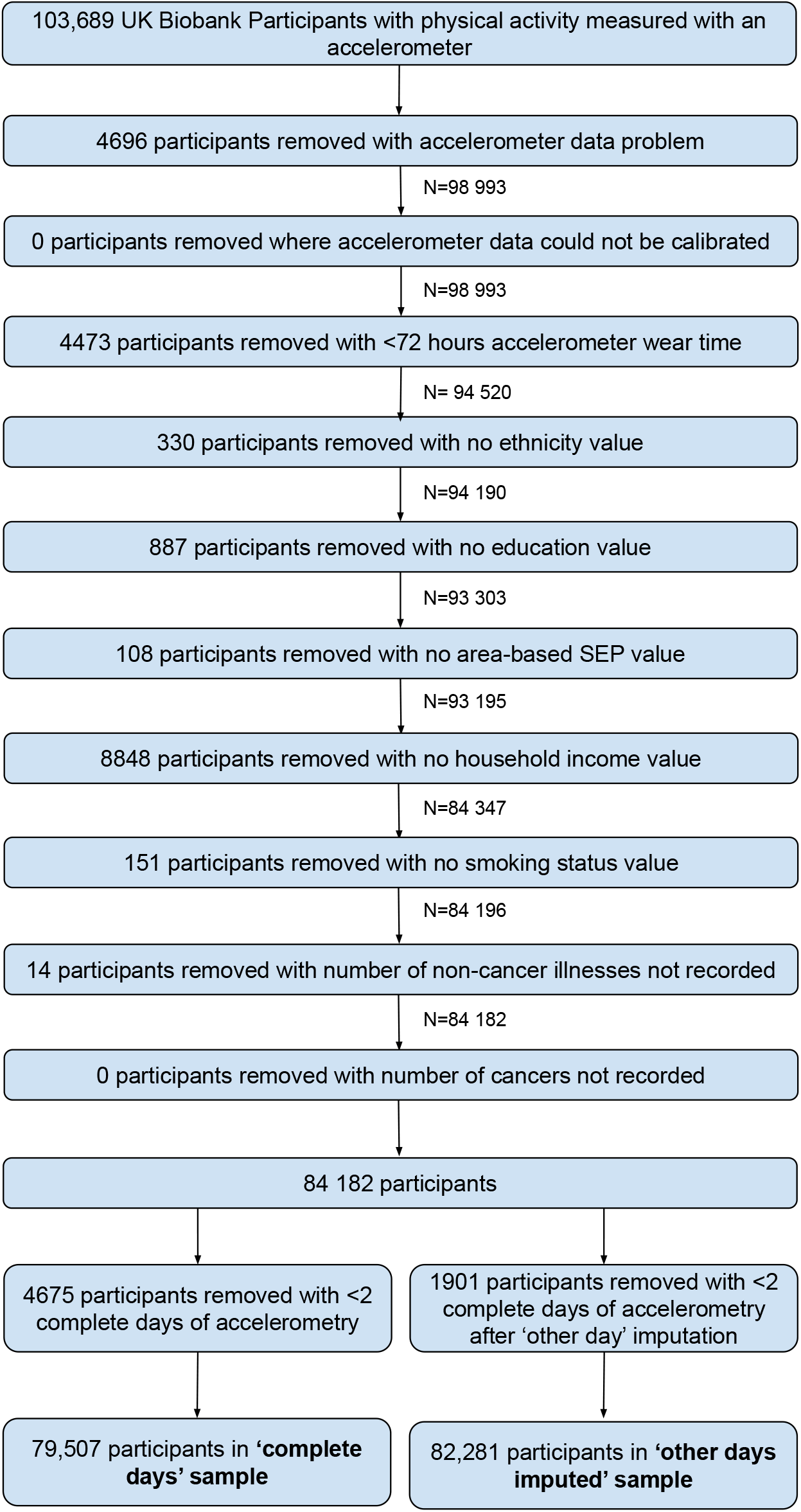
Participant flow diagram

### Data collection

#### Physical activity measurement

Physical activity was measured using the Axivity AX3 wrist-worn tri-axial accelerometer and stored in gravity (g) units. The devices were configured to start recording at 10am two working days after dispatching to participants by post, and to record tri-axial acceleration at a frequency of 100Hz (dynamic range +/- 8g) for 7 days. This results in over 181 million values per person (100 values per second across 7 days for each of the three axes).

#### All-cause mortality

Date of death was obtained from the NHS Information Centre for participants from England and Wales and the NHS Central Register for participants from Scotland.

#### Potential confounders

We considered the following to be likely confounding factors (based on their known or plausible effects on physical activity and mortality): sex, age, ethnicity, socio-economic position, smoking, BMI and general ill-health (Supplementary figure 3). Sex, ethnicity and smoking status (never, current or previous) were self-reported at the baseline assessment. We used education level, household total income and Townsend deprivation index (a score representing the deprivation of the participant’s neighbourhood) to reflect participant’s socio-economic position (Supplementary section S2 for details of these measures). At the baseline assessment weight was measured (to the nearest 100g) in light clothing and unshod using a Tanita BC418MA body composition analyser, and height to the nearest cm using a Seca 202 device. We used the number of baseline self-reported cancer and non-cancer illnesses as measures of baseline general ill-health.

The season in which participants wore the accelerometer, while not a confounder since it would not plausibly affect subsequent risk of death, may affect activity. We therefore included season (autumn [September-November], winter [December-February], summer [June-August] and spring [March-May]) as a covariate, to reduce the variation in activity exposure variables.

### Statistical analyses

#### Dealing with missing accelerometer data

While UK Biobank participants were asked to wear the accelerometer continuously for 7 days, 24% of our sample had some missing data. We identified periods of non-wear using the Biobank accelerometer analysis tool (available at https://github.com/activityMonitoring/), defined as consecutive stationary episodes (where all three axes had a standard deviation of less than 13.0 milli-gravities [m-grav]) lasting for at least 60 minutes (the sensitivity of the accelerometer makes it possible to detect very small movements indicating it is being worn) (17). We used two approaches to explore missing accelerometer data – a ‘complete days’ approach and an ‘other day’ imputation approach – that make different missingness assumptions. The complete days approach uses only days with complete accelerometer data in our analyses (referred to as ‘valid’ days). The other day imputation approach involved finding all other periods of accelerometer data on other days that are during the same time period and have no missing data (including from days with missing data at other times). One of these periods is then randomly chosen as the imputed sequence for the missing region. ‘Imputed valid’ days are those with no missing data after this imputation. Details on missing data assumptions of these approaches are provided in Supplementary section S3. We report results using the complete days data as our main results, and results using the imputed data are provided in the supplementary material.

#### Deriving physical activity bouts

We assigned each 1 minute epoch (interval) of accelerometer data to an activity category – either sleep, sedentary, light or MVPA. To do this we made use of a previously published machine learning model that predicts activity categories (sleep, sedentary, walking, light and MVPA) from accelerometer data, that was trained using accelerometer data captured in free-living conditions and labelled with ‘ground truth’ activities from accompanying videos and the Compendium of Physical Activities (18). We used a *hybrid* approach that first identified MVPA as minutes ≥100 m-grav (a threshold used in previous research (19)), and then used the machine learning model to identify minutes of sleep and sedentary behaviour from those not already assigned to MVPA (20). All other minutes not assigned to MVPA, sleep or sedentary categories were assigned to the light activity category. For each participant we identified contiguous sequences of 1-minute epochs with a given activity category; these are referred to as activity bouts and can be of any length so long as the participant remains in the same activity category.

As a sensitivity analysis we used only the machine learning model to define all categories, and refer to this as the *ML-only* approach. As well as categories of MVPA, sleep and sedentary this model predicts walking and light activity. There are two reasons we used the hybrid approach as our main analysis: 1) the degree of misclassification of the machine learning model for MVPA (e.g. only 58% of MVPA minutes were predicted correctly as MVPA (20)), and 2) the allocation of minutes to a separate walking category when brisk walking is considered to be moderate activity and slow walking would be light activity.

#### Deriving summary variables reflecting time spent in activity categories, overall and in bout length strata

For both approaches we calculated, for each participant, the overall time they spent in each activity category, on average per day. Hence, for each participant the total across all activity categories (for both the *hybrid* and *ML-only* approaches) equalled 1440, the number of minutes in a day. To investigate whether the association of time spent in MVPA (or sedentary) with all-cause mortality, changes depending on the time participants spend in bouts of different duration we categorised them as: short (1-15 minutes), medium (16-40 minutes) and long (41+ minutes). We derived the time spent in MVPA and sedentary bouts of each length, on average per day. Further details on our derivation of activity summary variables are provided in Supplementary section S4.

#### Estimating the association of overall time spent in activity categories with all-cause mortality taking account of total time spent in that activity and coupling of spending more time in one activity category with less time in another category

We used Cox proportional hazard regression to test the association of each activity summary variable with all-cause mortality. All models were performed with age as the time variable. For each participant follow-up was defined as starting at the age they stopped wearing the accelerometer. End of follow up was defined as either participant’s age on 30^th^ November 2016 for those in Scotland and 31^st^ January 2018 for those in England and Wales (the last day of the last month for which the number of deaths was greater than 90% of the mean of the preceding three months), or their age at death for those who died before the end of follow-up. We tested each association before and after adjustment for potential confounders. Exact dates of birth were not available so ages were estimated assuming birth on 1^st^ July in the reported year of birth.

It is possible that BMI and ill-health subsequent to activity assessment could mediate the effect of activity on mortality. Whilst we adjusted for BMI and ill-health assessed at baseline (3-9 years prior to activity assessment), tracking of these factors across time (e.g. due to factors that affect BMI across the life-course) means that BMI and ill-health measured before activity are also proxies for these factors measured after activity (Supplementary figure 2). Adjusting for proxies of mediating factors could attenuate our estimates towards the null (21). We therefore performed a sensitivity analysis excluding BMI and ill-health as covariates.

Within 1 day there are 1440 minutes so a greater amount of time spent in one activity category must be coupled with a lesser amount of time spent in one or more other activity categories. For this reason we model associations in terms of couplings of activity categories, in a similar way to our previous activity bigrams approach (22). We assign, in turn, one activity category as the baseline and estimate the hazard associated with spending 10 minutes less time in this baseline category when coupled with spending 10 minutes more time in a given comparison category, on average per day. Further details of this approach are provided in Supplementary section S5.

#### Relating time spent in short, medium and long bouts of MVPA or sedentary with all-cause mortality

As with our models for overall time, to estimate the association of time spent in sedentary and MVPA bouts of different length with all-cause mortality, we model these associations in terms of couplings of activity categories. Hence, unlike previous work (6-8,14), our models estimate associations for couplings between MVPA and sedentary bouts and also with other activity categories. Further details are provided in Supplementary section S6.

For all models we generated Schoenfeld residuals and estimated the correlation between log-transformed survival time and the scaled Schoenfeld residuals to test the proportional hazards assumption.

Analyses were performed in R version 3.5.1, Matlab r2015a or Stata version 15, and all of our analysis code is available at [https://github.com/MRCIEU/UKBActivityBoutLength/]. Git tag v0.1 corresponds to the version of the analyses presented here.

## RESULTS

Of the 84,182 eligible participants 79,507 and 82,281 were included in our complete days and other day imputed samples, respectively (Figure 1). Other day imputation greatly increased the number of valid days (e.g. 96% and 24% of participants had 7 valid days in the imputed and complete days data; Supplementary figure 3). The total person years at risk in the complete days analysis was 249,592 and 761 participants died, giving a mortality rate of 3.05 per 1000 person years (equivalent numbers for other day imputed sample were 258,263 and 796, with a mortality rate of 3.08 per 1000 person years). Participants who were included in our analyses, compared with those who were invited to wear an accelerometer but did not respond, did not accept or had missing accelerometer of confounder data, were younger, more likely to be white, more educated, and living in an area with less social deprivation, had a higher income, lower BMI, were less likely to have ever smoked, had fewer illnesses, were more likely to have worn the accelerometer in winter and were less likely to die (Supplementary table 1).

Supplementary table 2 shows the distributions of the average number of minutes per day in each activity category. Sedentary time was more often accumulated in bouts of longer duration, and MVPA was more often accumulated in shorter bouts. Participants with more time sedentary on average spent more time in long sedentary bouts and less time in short or medium sedentary bouts (Supplementary table 3). Participants who spent more time in MVPA on average spent more time in all categories of MVPA bout length, but particularly shorter bouts. On average, participants spent more time in MVPA using the hybrid approach compared with the ML-only approach. Patterns of correlation of overall time sedentary or in MVPA with bout length categories were similar for the hybrid and ML-only approaches. Correlations between the same characteristics derived using the hybrid and ML-only approaches were variable. Overall time sedentary was very strongly correlated (Pearson’s rho > 0.99), with reasonable correlation for the sedentary bout length categories (Pearson’s rho’s of 0.73, 0.76 and 0.96 for short, medium and long sedentary bouts, respectively), and lower correlations for MVPA (e.g. Pearson’s rho = 0.43 for overall time spent in MVPA and 0.26 for long MVPA bouts).

### Associations of overall time spent in activity categories, with all-cause mortality

Associations of time spent in activity categories are shown in Figure 2. Overall, time spent in the different activity categories relates differently to mortality. Spending more time in MVPA was associated with lower mortality when coupled with less time spent sleeping, sedentary or in light activity, and these associations were of a similar magnitude (e.g. HR 0.93 [95% CI: 0.91, 0.95] and 0.95 [95% CI: 0.92, 0.97] for 10 minutes more MVPA coupled with 10 minutes less sedentary and light activity, respectively). Spending more time in light activity was also associated with lower mortality when coupled with less time spent sleeping or sedentary (HRs 0.99 [95% CI: 0.97, 1.00] and 0.98 [95% CI: 0.97, 0.99], respectively). Results of sensitivity analyses using the ML-only approach were largely consistent, although there were some differences (e.g. spending more time in light activity coupled with less time sleeping or sedentary were consistent with the null; Supplementary table 4 and Supplementary figure 5).

**Figure 2:**
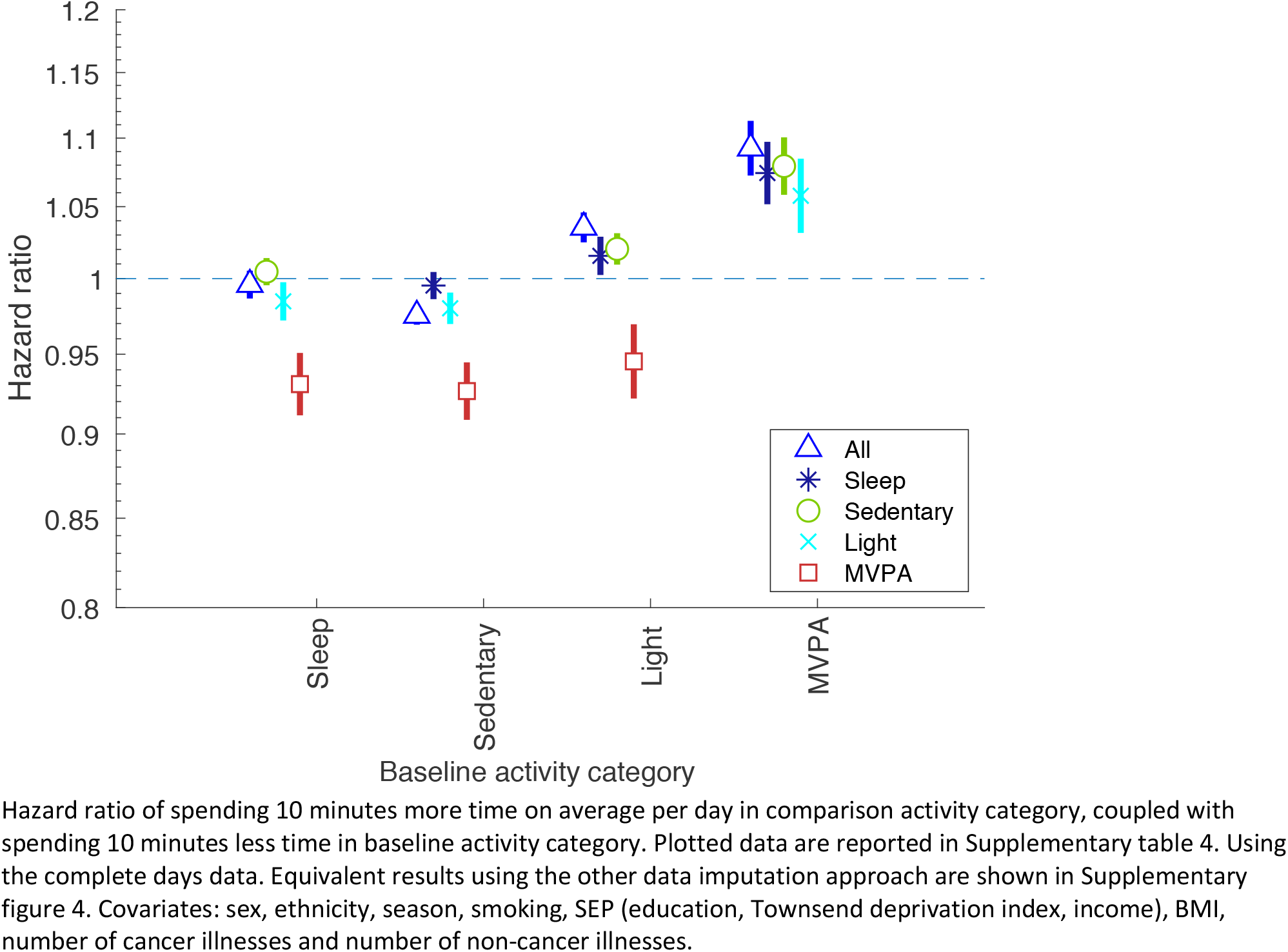
Associations of less time spent in baseline activity category coupled with more time in comparison category, with all-cause mortality

### Associations of MVPA and sedentary bout length with all-cause mortality

We found little evidence to suggest that associations differed across MVPA bout lengths (Figure 3a and Supplementary table 5). For example, our estimate of association for spending 10 minutes less time in short MVPA bouts (<16 minutes duration) coupled with spending 10 minutes more time in long MVPA bouts (40+ minutes duration), with all-cause mortality, was consistent with the null (HR 1.05 [95% CI: 0.93, 1.19]). We also found little evidence that associations differed across sedentary bout lengths (Figure 3b and Supplementary table 6). For example, our estimate of association for spending 10 minutes less time in short sedentary bouts coupled with spending 10 minutes more time in long sedentary bouts, with all-cause mortality, was consistent with the null (HR 1.04 [95% CI: 0.99, 1.09]). Sensitivity analyses using the ML-only approach showed some differences compared with the hybrid approach (Supplementary figure 7). Most notably, they suggest that spending less time in shorter sedentary bouts coupled with spending more time in longer sedentary bouts, associates with a lower all-cause mortality.

**Figure 3:**
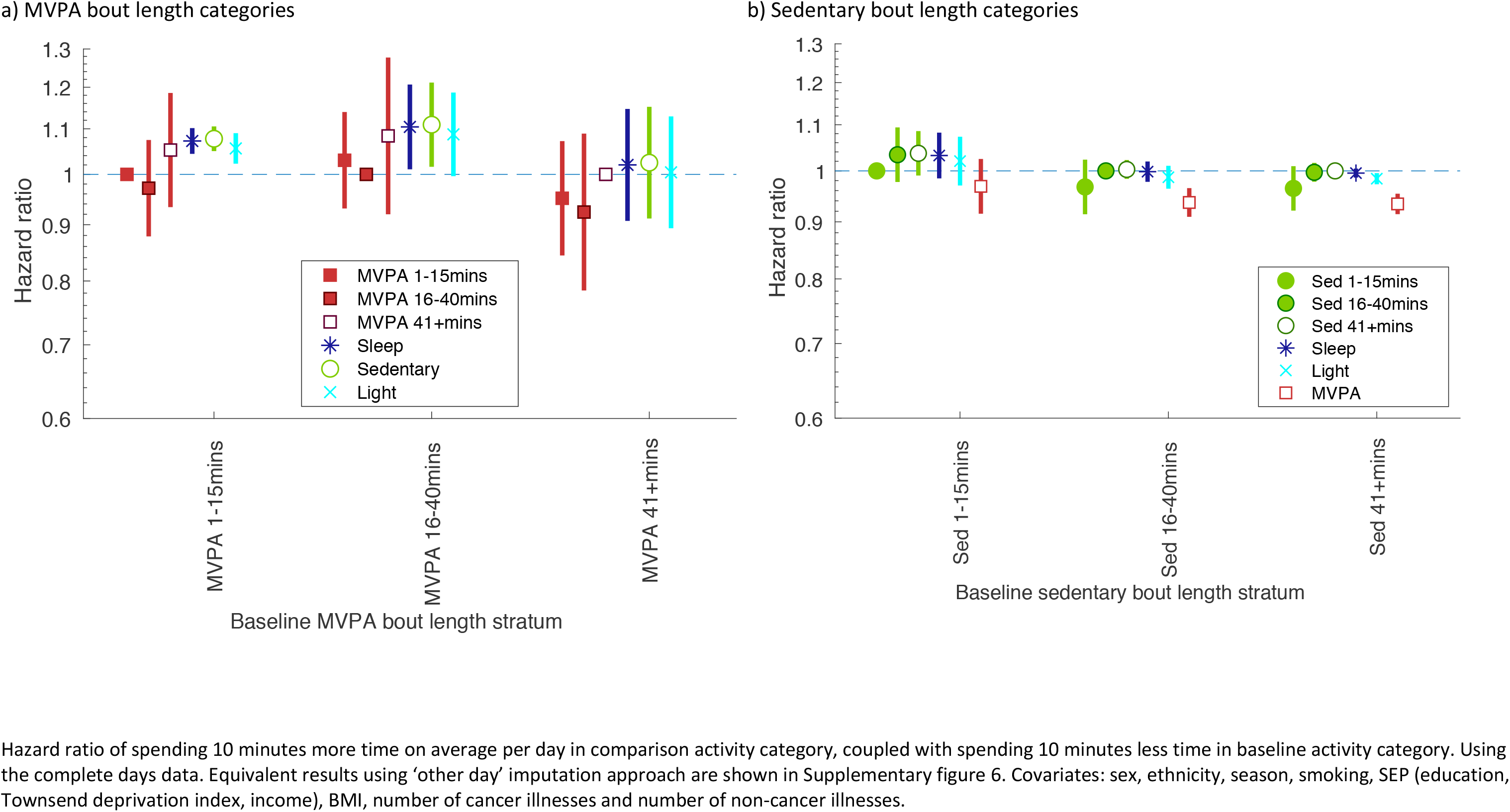
Associations of time spent in MVPA and sedentary bouts of given duration, with all-cause mortality

Results of sensitivity analyses using ‘other day’ imputed data were broadly consistent with the results of our main analyses using the complete days data (Supplementary figures 5-7, and Supplementary tables 4-6). Results of sensitivity analyses excluding BMI and ill-health as covariates were comparable to our main analyses (Supplementary tables 4-6).

We found little evidence of violation of the proportional hazards assumption across all Cox regression models (Supplementary tables 4-6).

## DISCUSSION

We have uniquely shown that time spent in MVPA is associated with lower mortality, irrespective of whether it is coupled with less time spent sleeping, sedentary or in light activity, and irrespective of whether it is obtained from several short bouts or fewer longer bouts. We have also shown that time spent sedentary is associated with a higher mortality if it is coupled with less time in light activity (but to a lesser extent than when coupling with MVPA). These findings emphasize the specific importance of MVPA. They also support recent policy changes in some countries that have removed the suggestion that MVPA should be accumulated in bouts of 10 minutes per day. Those policy changes were made on the basis of randomized controlled trial evidence but the trials were small, only looked at associations with continuously measured risk factors, and were deemed to have high risk of bias. Our results do not support the specific promotion of accumulating MVPA in several smaller bouts but rather suggest accumulating MVPA in any bout length could reduce risk of premature mortality, but also that replacing sedentary periods of any length with light activity could be beneficial, although to a lesser extent. This is an important public health message as it allows people with different preferences and lifestyles to improve health through accumulating activity in different ways.

Importantly, the methods that we have used here address limitations of other studies that appear not to have controlled for overall time spent across all bout lengths of a given activity category (14), considered that greater amounts of one activity should be coupled with lesser amounts of another (7,8,14), or assessed each coupling combination (6–8,14). We provide all of our code [https://github.com/MRCIEU/UKBActivityBoutLength/] so that others can use this method for exploring other outcomes, or risk factors for different patterns of activity, and examine associations in other populations.

There are no previous studies assessing the association of MVPA bout length with mortality. Our findings contrast with those of a previous study that analysed sedentary bout length in terms of the percentage of sedentary time in each sedentary bout length strata and concluded that longer versus shorter sedentary bouts were associated with a higher risk of premature mortality (14). We hypothesise that their results may be explained by an effect of total time spent sedentary on all-cause mortality, which was not taken into account in that study.

### Study strengths and limitations

Strengths of the study include the large sample size and use of accelerometer data rather than self-report to measure activity, and the prospective nature of the study. We have developed and used a method that appropriately accounts for coupling of activities. We have appropriately explored associations of total time spent in MVPA and sedentary with mortality, including whether this differs by bout length and depending on what alternative activity it is coupled with. We undertook sensitivity analyses to assess missing data assumptions. The code for generating our variables is freely available so can be used by others to explore associations with other health outcomes in UK Biobank and in other studies with similar activity data.

Our study has a number of limitations. We used a previously published machine learning model to predict activity categories and so it is possible that misclassifications of those predictions biased our estimates of association. However, our main analysis used a hybrid approach where MVPA was identified using a threshold (>100 m-grav), since prediction accuracy for MVPA from the machine learning model was particularly low. We also conducted sensitivity analyses using the machine learning predictions only (ML-only). These results were largely consistent for associations with overall time spent in each activity category, but showed some differences for our bout length results that may be due to biases in the types of activities assigned as MVPA by the ML-only approach compared with the hybrid approach.

Participants tended to spend relatively little time in MVPA (particularly in long bouts) so these estimates were imprecise. Further studies are needed in larger samples (e.g. when larger cohort studies are created) and with more precise measures of MVPA activity bouts (e.g. through more accurate prediction of MVPA using machine learning) to further explore these associations.

While we accounted for known, measured confounders, our analyses may be biased by residual confounding. For example, it is possible that dietary factors and access to green space or facilities to be physically active, might confound our associations. Adjustment for three different measures of socioeconomic position, including an area-based measure and BMI, are likely to have controlled for some of the potential confounding by these. Sensitivity analyses restricting follow-up to those alive 5 years after wearing the accelerometer would help to ensure associations are not due to confounding via existing ill-health, but follow-up periods in UK Biobank are currently too short to be able to do this.

UK Biobank is a highly selected sample of the UK population with a response rate of 5.5% (23), and evidence suggests that those who volunteered are a more affluent and healthy population than those who did not (15). The participants who were included here were also a more affluent and healthier group than UK biobank participants who were not included. This “selection” may mean our estimates are biased (Supplementary section S7 for further details). Most of the participants in UK Biobank are of White European origin and our results may not generalise to other populations.

To conclude, we have used a novel approach to assess whether time spent in different activity types, and in short, medium or long bouts of MVPA and sedentary behaviour, are associated with all-cause mortality. Our study confirms a strong association between active time and reduced mortality, and the extra benefit of MVPA compared with light activity. We found little evidence that associations with time spent in MVPA or sedentary differ according to bout length. These results support the recent decision to amend the UK and US physical activity guidelines to remove the advice that MVPA should be accumulated in bouts of 10 minutes or more (3,4). Further work is needed to replicate our results in independent data and to investigate causality. Finally, our results highlight the importance of the isotemporal ‘coupling of time’ perspective and suggest that this should be commonplace in any activity analyses, as public health advice based on increasing time spent in a given activity type is misleading without accompanying details of the activities from which this time should be taken.

## Data Availability

The data underlying the results presented in the study are available from UK Biobank [www.ukbiobank.ac.uk].

## ACKNOWLEDGEMENTS

This research has been conducted using the UK Biobank Resource under Application Number 17810.

## AUTHOR CONTRIBUTIONS

LACM conceptualized the study, conceived the activity bouts approach, contributed to the design of the study, performed all analyses, wrote the first version of the manuscript, critically reviewed and revised the manuscript and approved the final version of the manuscript as submitted. DAL contributed to the design of the study, critically reviewed and revised the manuscript and approved the final version of the manuscript as submitted. KT contributed to the design of the study, critically reviewed and revised the manuscript and approved the final version of the manuscript as submitted. TRG contributed to the design of the study, critically reviewed and revised the manuscript and approved the final version of the manuscript as submitted. DC contributed to the design of the study, critically reviewed and revised the manuscript and approved the final version of the manuscript as submitted.

